# Estimating the cost of antibiotic use on future collateral resistance: a retrospective comparison of cefuroxime versus cefazolin and amoxicillin/clavulanate

**DOI:** 10.1101/2022.01.10.22269003

**Authors:** Michal Chowers, Tamir Zehavi, Bat-Sheva Gottesman, Avi Baraz, Daniel Nevo, Uri Obolski

**Author notes:** Corresponding author: Uri Obolski; Mail, School of Public Health and the Porter School of the Environment and Earth Sciences, Tel Aviv University, Ramat Aviv, Tel Aviv, Israel, 6997801. Alternate corresponding author: Michal Chowers; Mail, Infectious diseases unit, Meir medical center, 59 Tsharnichovski st, Kfar Saba, Israel 44281.

## Abstract

**Background:** Quantitative estimates of collateral resistance induced by antibiotic use are scarce. This study compared the effects of treatment with amoxicillin/clavulanate or cefazolin, compared to cefuroxime, on future resistance to ceftazidime among hospitalized patients.

**Methods:** A retrospective analysis of patients with positive bacterial cultures hospitalized in an Israeli hospital during 2016-2019 was conducted. Patients were restricted to those treated with either amoxicillin/clavulanate, cefazolin, or cefuroxime and re-hospitalized with a positive bacterial culture during the following year. A 1:1 matching was performed for each patient in the amoxicillin/clavulanate and cefazolin groups, to a single patient from the cefuroxime group, yielding 185:185 and 298:298 matched patients. Logistic regression and g-formula (standardization) were used to estimate the odds ratio (OR), risk difference (RD), and number needed to harm (NNH).

**Results:** Cefuroxime induced significantly higher resistance to ceftazidime than amoxicillin/clavulanate or cefazolin: the marginal OR was 1.76)95%CI 1.16-2.83) compared to amoxicillin/clavulanate, and 1.98 (95%CI 1.41-2.8) compared to cefazolin; The RD was 0.118 (95%CI 0.031-0.215) compared to amoxicillin/clavulanate, and 0.131 (95%CI 0.058-0.197) compared to cefazolin. We also estimated the NNH: replacing amoxicillin/clavulanate or cefazolin with cefuroxime would yield ceftazidime-resistance in one more patient for every 8.5 (95% CI 4.66-32.14) or 7.6 (95% CI 5.1-17.3) patients re-hospitalized in the following year.

**Conclusions:** Our results indicate that treatment with amoxicillin/clavulanate or cefazolin is preferable to cefuroxime, in terms of future collateral resistance. The results presented here are a first step towards quantitative estimations of the ecological damage caused by different antibiotics.

**Key points:** We performed a retrospective study estimating collateral resistance of treatment with cefuroxime relative to amoxicillin/clavulanate or cefazolin. Application of novel analytical methods allowed us to estimate the number needed to harm and hence ecological damage of the different treatments.

## Introduction

Antibiotic resistance is a growing problem worldwide [1, 2]. Although it is clear that increased antibiotic use drives resistance, the quantity and breadth of antibiotic use continues to rise [3]. This is driven by advances in medical interventions, increasing numbers of immunocompromised individuals and aging of the population. Moreover, increased resistance itself is a driving factor of broad-spectrum antibiotic use.

Appropriate antibiotic usage includes incorporating the knowledge of the expected bacterial culprit and local antibiogram, as well as the spectrum of the antibiotic considered, its ability to induce resistance and its cost. While the spectrum and cost of antibiotics are clear, their ability to induce resistance is hard to measure accurately. For example, it has been shown that quinolones and macrolides can readily induce resistance, but this effect is not easily measurable [4-7]. Another well-known observation is that cephalosporins induce more resistance than penicillin [8]. Moreover, attempts have been made to rank the ecological consequences of some beta-lactam antibiotics using expert consensus [9]. However, a quantitative measure, enabling comparison between specific drugs, such as broad-spectrum penicillins to narrower spectrum cephalosporins, or quinolones to cephalosporins, is lacking. This measure could facilitate informed decisions of the treating physician, and hopefully lower future resistance.

The study aim was to examine the effect of cefuroxime versus amoxicillin/clavulanate or cefazolin, on the subsequent occurrence of resistance among hospitalized patients. These antibiotics were chosen due to their wide use in the study hospital. Resistance to ceftazidime was selected as the outcome variable, because it serves as a reliable marker for extended spectrum beta-lactamase (ESBL) resistance. [10]

## Methods

Study design: This historical cohort study was based in Meir Medical Center, a secondary 740-bed university-affiliated hospital located in central Israel. It serves a population of about 600,000 individuals, with a similar number of admissions per year.

### Participants

Our dataset included all adult patients with a positive bacterial culture in Meir Medical Center from January 1, 2016, to December 31, 2019. Patients who received either cefazolin, cefuroxime or amoxicillin-clavulanate for the first recorded time, termed the ‘baseline’ hospitalization, were included. Patients were further restricted to those who had a subsequent hospitalization with a culture tested for ceftazidime resistance, within a year from the baseline hospitalization. Excluded were patients who received any other antibiotic (except metronidazole, trimethoprim-sulfamethoxazole or penicillin) during their baseline hospitalization or within the one-year period.

#### Data

Variables collected from patient files included age, sex, comorbidities, antibiotic treatment, days from baseline admission to culture, duration of baseline admission and culture source. If available, we collected culture results and culture source from the baseline hospitalization in which antibiotics were given.

#### Ethics

The study was approved by the Institutional Review Board (Helsinki) Committee of Meir Medical Center. Since this was a retrospective study, using archived medical records, an exemption from informed consent was granted by the Helsinki Committee.

### Statistical analysis

#### Matching

The treatment groups compared were patients who received either cefazolin or amoxicillin-clavulanate against patients who received cefuroxime. To achieve balance between the treatment groups, we performed 1:1 matching without replacement. Each patient in the cefazolin and amoxicillin/clavulanate groups was matched to one patient from the cefuroxime group. First, patients were exactly matched based on the existence and result of a “baseline” culture obtained prior to the use of the antibiotic in question. That is, patients with a ceftazidime resistant, susceptible or no culture taken were matched between the two treatment groups. Then, we examined several matching metrics, including Mahalanobis distance, and propensity score (PS) matching with a caliper. The PS for receiving each treatment was calculated using a multivariate logistic regression model detailed below. Importantly, matching can be performed in any way desired and until balance is achieved – as long as it is assessed by the covariates’ balance without the outcome variable [11].

Balance of patient covariates was assessed by inspecting post-matching tables of patient covariates and PS distributions. P-values in the tables were calculated using a t-test for continuous variables, and a chi-square test for categorical variables. P-values were only used as indication of substantial differences between the groups and not for inference, and as such were not corrected for multiple comparisons.

For both treatment comparisons, optimal balance was achieved by a combination of Mahalanobis distance and a caliper on the PS.

When comparing amoxicillin/clavulanate with cefuroxime, the optimal balance was achieved by matching based on the Mahalanobis distance applied to the variables log(age), log(days of initial admission), sex, diabetes, and chronic obstructive pulmonary disease (COPD). In addition, a caliper of 0.5 standard deviations was applied to the PS, where the PS model included, log(age), log(days of initial admission), and indicators of dementia, immunosuppression and COPD. One patient in the amoxicillin/clavulanate treatment group could not be adequately matched and was dropped from further analyses.

When comparing cefazolin with cefuroxime, the best balance was achieved by matching based on the Mahalanobis distance applied on the variables age, days of initial admission, and diabetes. In addition, a caliper of 0.45 standard deviations was applied to the PS, where the PS model included age, days of initial admission, and sample source. One patient in the cefazolin treatment group could not be adequately matched and was dropped from further analyses.

To facilitate balance assessment, another PS model including all the variables used in the matching process was calculated for each treatment pair. The final results of the matching process are presented in Figure 2 and in Tables 2-3.

#### Estimation

After matching, several measures of association were computed between the matched groups and the outcome.

The odds ratios (OR) estimates were calculated by including all the variables employed in the matching process in a multivariate logistic regression model with resistance as the outcome. Then, we calculated the marginal ORs using the g-formula: the logistic regression models were used to estimates the probability of resistance when assuming a) that all patients received the non-cefuroxime antibiotic and then b) that all patients received cefuroxime. The estimated results were averaged and then transformed to the OR. To obtain confidence intervals (CI) for these point estimates, we employed the BCa bootstrap CI with 999 repetitions, at the matched pairs level [12].

To verify our model’s robustness, we applied another approach - univariate logistic regression on the matched data with treatment as the only independent variable and resistance to ceftazidime as the outcome. The CIs for these ORs were estimated using cluster-robust standard errors, setting the clusters as the matched pairs [12].

Risk difference (RD) was also estimated using the g-formula, by an analogous approach to the abovementioned OR estimation, with the only difference being that the predicted probabilities were subtracted rather than transformed to ORs [13].

All analyses were performed in R, using the *Matching* [14], *boot* [15], and *sandwich* [16] packages.

## Results

The resulting datasets included 186, 299, and 857 patients treated with amoxicillin/clavulanate, cefazolin, and cefuroxime, respectively (Figure 1).

**Figure 1:**
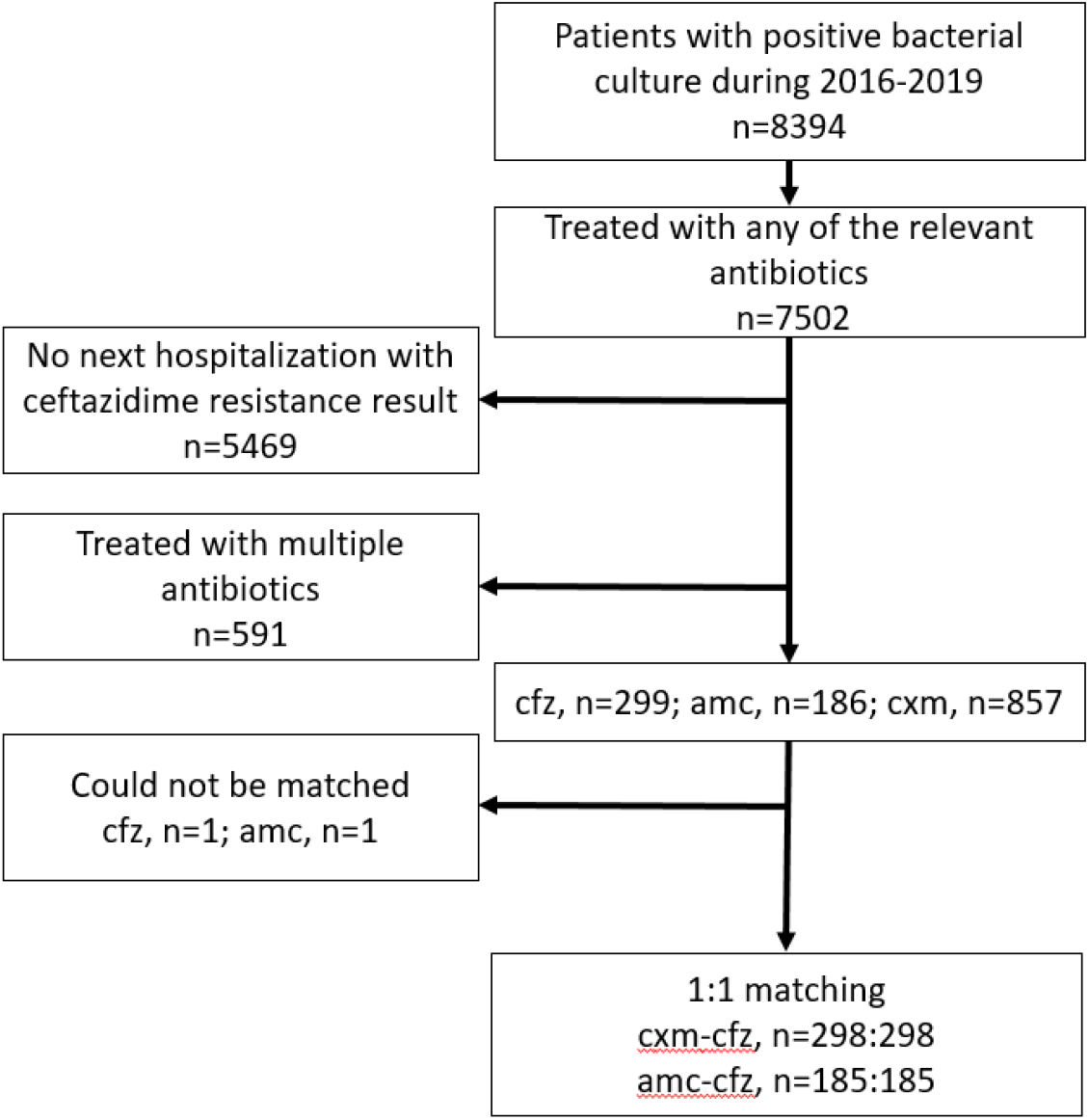
Flowchart of study participants. Abbreviations: amc=amoxicillin/clavulanate, cfz=cefazolin, cxm=cefuroxime.

The characteristics of patients in the treatment groups are presented in Table 1. Since the characteristics differed significantly, a 1:1 matching was performed. The cefuroxime treatment group was always larger, as it is a common first line treatment at Meir Medical Center. Hence, it served as the ‘treatment’ (rather than ‘control’) group. That is, we matched a single patient from the cefuroxime group to each of the patients in the non-cefuroxime groups.

**Table 1:**
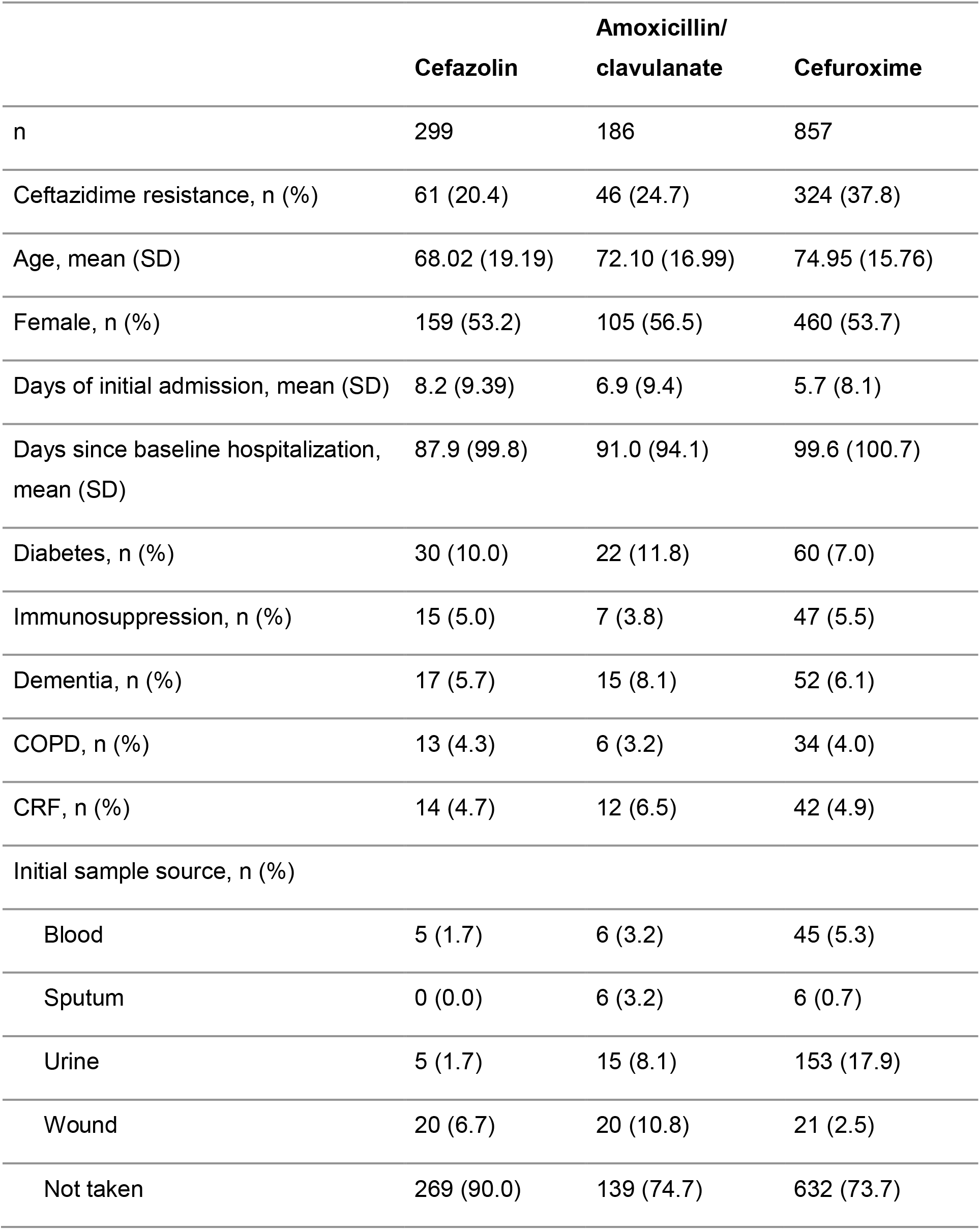
Patient covariates for all three treatments.

|  | **Cefazolin** | **Amoxicillin/**<br>**clavulanate** | **Cefuroxime** |
| --- | --- | --- | --- |
| n | 299 | 186 | 857 |
| Ceftazidime resistance, n (%) | 61 (20.4) | 46 (24.7) | 324 (37.8) |
| Age, mean (SD) | 68.02 (19.19) | 72.10 (16.99) | 74.95 (15.76) |
| Female, n (%) | 159 (53.2) | 105 (56.5) | 460 (53.7) |
| Days of initial admission, mean (SD) | 8.2 (9.39) | 6.9 (9.4) | 5.7 (8.1) |
| Days since baseline hospitalization,<br>mean (SD) | 87.9 (99.8) | 91.0 (94.1) | 99.6 (100.7) |
| Diabetes, n (%) | 30 (10.0) | 22 (11.8) | 60 (7.0) |
| Immunosuppression, n (%) | 15 (5.0) | 7 (3.8) | 47 (5.5) |
| Dementia, n (%) | 17 (5.7) | 15 (8.1) | 52 (6.1) |
| COPD, n (%) | 13 (4.3) | 6 (3.2) | 34 (4.0) |
| CRF, n (%) | 14 (4.7) | 12 (6.5) | 42 (4.9) |
| Initial sample source, n (%) |  |  |  |
| Blood | 5 (1.7) | 6 (3.2) | 45 (5.3) |
| Sputum | 0 (0.0) | 6 (3.2) | 6 (0.7) |
| Urine | 5 (1.7) | 15 (8.1) | 153 (17.9) |
| Wound | 20 (6.7) | 20 (10.8) | 21 (2.5) |
| Not taken | 269 (90.0) | 139 (74.7) | 632 (73.7) |

Appropriate balance between the groups was achieved, as can be seen in plots of the PS distributions (Figure 2) and in patients’ characteristics after matching (Tables 2, 3). We note that the achieved balance was sufficient even for variables not used in the matching process (see Methods for variables used). Importantly, we could not adjust for the days elapsed from the first treatment to the outcome culture, as it is a post-treatment variable and doing so might induce bias. Nonetheless, this variable was well-balanced after matching — indicating a similar patient population was successfully matched. This matching process emulated a conditionally randomized trial among patients who typically received cefazolin or amoxicillin/clavulanate and would instead have received cefuroxime (analogous to estimation of the average treatment effect on the untreated, ATU) [17, 18].

**Figure 2:**
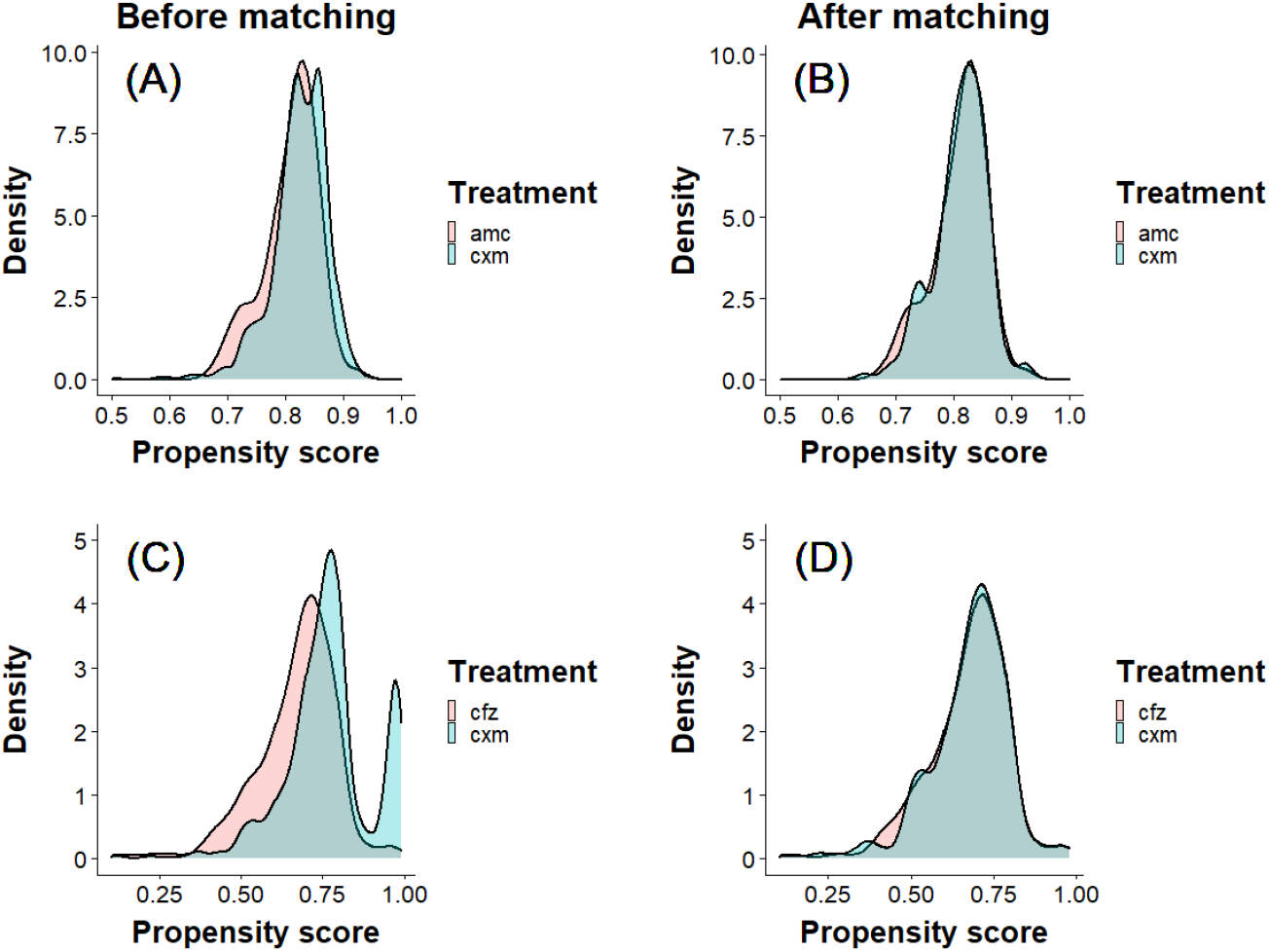
Propensity score distributions before and after matching. The propensity scores of amoxicillin/clavulanate (A, B; red) and cefazolin (C, D; red) are plotted in red. Overlain are the propensity score distributions of cefuroxime before (A, C; blue) and after (B, D; blue) matching. amc=amoxicillin/clavulanate, cfz=cefazolin, cxm=cefuroxime.

**Table 2:** Patient covariates after 1:1 matching of patients treated with cefuroxime to those treated with cefazolin. SMD= standardized mean difference.

|  | **Cefazolin** | **Cefuroxime** | **SMD** | **p-value** |
| --- | --- | --- | --- | --- |
| n | 298 | 298 |  |  |
| Age, mean (SD) | 68.17 (19.05) | 68.72 (18.50) | -0.029 | 0.717 |
| Female, n (%) | 158 (53.0) | 158 (53.0) | 0.000 | 1 |
| Days of initial admission, mean (SD) | 8.07 (9.2) | 8.03 (9.5) | 0.004 | 0.958 |
| Days since baseline hospitalization, mean (SD) | 87.95 (99) | 89.74 (96.1) | -0.018 | 0.82 |
| Diabetes, n (%) | 30 (10.1) | 28 (9.4) | 0.023 | 0.89 |
| Immunosuppression, n (%) | 15 (5.0) | 11 (3.7) | 0.060 | 0.55 |
| Dementia, n (%) | 17 (5.7) | 15 (5.0) | 0.030 | 0.86 |
| COPD, n (%) | 13 (4.4) | 9 (3.0) | 0.068 | 0.52 |
| Chronic renal failure, n (%) | 14 (4.7) | 11 (3.7) | 0.047 | 0.68 |
| Initial sample source, n (%) |  |  |  | 0.08 |
| Blood | 5 (1.7) | 9 (3.0) | -0.101 |  |
| Sputum | 0 (0.0) | 2 (0.7) | NA |  |
| Urine | 5 (1.7) | 5 (1.7) | 0.000 |  |
| Wound | 20 (6.7) | 8 (2.7) | 0.160 |  |
| Not taken | 268 (89.9) | 274 (91.0) | -0.066 |  |

**Table 3:** Patient covariates after 1:1 matching of patients treated with cefuroxime to those treated with amoxicillin/clavulanate. SMD= standardized mean difference.

| **Patient covariates** | **Amoxicillin/**<br>**clavulanate** | **Cefuroxime** | **SMD** | **p-value** |
| --- | --- | --- | --- | --- |
| n | 185 | 185 |  |  |
| Age, mean (SD) | 72.05 (17) | 73.09 (16.2) | -0.061 | 0.546 |
| Female, n (%) | 105 (56.8) | 105 (56.8) | 0.000 | 1 |
| Days of initial admission, mean (SD) | 6.74 (9.2) | 6.49 (9.2) | 0.027 | 0.788 |
| Days since baseline hospitalization, mean (SD) | 91.42 (94.1) | 99.09 (99.7) | -0.082 | 0.447 |
| Diabetes, n (%) | 22 (11.9) | 20 (10.8) | 0.034 | 0.87 |
| Immunosuppression, n (%) | 7 (3.8) | 4 (2.2) | 0.084 | 0.54 |
| Dementia, n (%) | 14 (7.6) | 10 (5.4) | 0.083 | 0.527 |
| COPD, n (%) | 6 (3.2) | 4 (2.2) | 0.057 | 0.749 |
| Chronic renal failure, n (%) | 12 (6.5) | 13 (7.0) | -0.020 | 1 |
| Initial sample source, n (%) |  |  |  | 0.01 |
| Blood | 6 (3.2) | 9 (4.9) | -0.097 |  |
| Sputum | 5 (2.7) | 1 (0.5) | 0.136 |  |
| Urine | 15 (8.1) | 24 (13.0) | -0.180 |  |
| Wound | 20 (10.8) | 6 (3.2) | 0.245 |  |
| Not taken | 139 (75.1) | 145 (78.4) | -0.076 |  |

Next, we estimated the marginal OR for ceftazidime resistance in the antibiotic treatment groups by applying a g-formula with a logistic regression model that included all the variables used for matching (see Methods). The marginal OR of cefuroxime treatment compared to amoxicillin/clavulanate was 1.76, 95%CI (1.16, 2.83) and 1.98 95%CI (1.41, 2.8) compared to cefazolin.

The estimates were robust and remained very similar when employing a different estimation method, using univariate logistic regression (after matching): amoxicillin/clavulanate (OR 1.81, 95%CI 1.14-2.88), and cefazolin (OR 1.90, 95%CI 1.33-2.73).

Using the g-formula, we were able to estimate RD for resistance to ceftazidime in addition to the OR. Similarly, for the outcomes in the OR, the RD analysis revealed that cefuroxime induced significantly higher resistance to ceftazidime than amoxicillin/clavulanate (RD 0.118, 95%CI 0.031-0.215) or cefazolin (RD 0.131, 95%CI 0.058-0.197).

Finally, the RD estimates allowed for estimation of the number needed to harm (NNH). We estimated that for every 8.5 (95% CI 4.66-32.14) patients treated with cefuroxime instead of amoxicillin/clavulanate, an additional ceftazidime-resistant infection would occur in one patient among those hospitalized with a bacterial infection in the following year. Likewise, for every 7.6 (95% CI 5.1-17.3) patients treated with cefuroxime instead of cefazolin, an additional ceftazidime-resistant infection would occur in one patient among those hospitalized with a bacterial infection in the following year.

## Discussion

The results of this study are noteworthy in two aspects. First, is the application of causal statistical methods to investigate antibiotic resistance. These methods enabled us to estimate the risk difference and therefore the NNH, in the context of future resistance, of different treatments [13]. Second, are the results of a comparison between a second-generation cephalosporin to either amoxicillin/clavulanate or to a first-generation cephalosporin, in terms of their ability to induce collateral resistance. We estimated that if treatment with amoxicillin/clavulanate or cefazolin would have been replaced with cefuroxime, one additional patient for every 8.5 (95% CI 4.66-32.14) or 7.6 (95% CI 5.1-17.3) treated patients hospitalized in the following year, would have a ceftazidime resistant, rather than sensitive, infection.

Causal relationships can be directly estimated from randomized controlled studies, but can also be approximated, under certain assumptions, from observational studies. In order to imply causality in our study, we employed matching, coupled with the g-formula (also known as standardization) to obtain a marginal OR, RD and NNH estimates. The validity of these causal inference methods requires three assumptions, all of which were reasonably fulfilled in our current study: 1. Overlap of patients with similar characteristics between treatment groups (also known as positivity), 2. Confounding was fully adjusted for, and 3. No interference between patients and no multiple versions of the treatment leading to different outcomes [17]. Thus, we estimated the treatment effect by comparing ceftazidime resistance in a scenario in which all patients in the matched cohort received amoxicillin-clavulanate or cefazolin, versus a scenario in which all patients received cefuroxime. The difference between the two options allowed for the marginal OR, RD and NNH estimates [18].

Clinical guidelines present several antibiotic treatment options with similar recommendations for different clinical scenarios [19, 20]. The decision on a specific antibiotic regimen relies on local antibiograms, cost, safety profile and ease of administration. Another important factor to consider is the influence of current antibiotic treatment on future resistance [21]. Yet, this effect is not easily quantifiable. The existing data rely mainly on case-control studies assessing risk-factors for resistance, thereby providing odds ratios for resistance. In contrast, the method employed here also provides risk for future resistance as a NNH, which is easily understood and intuitive for the practicing physician. Moreover, the NNH can be used to compare the ecological cost of different antibiotics and hence, can be taken into consideration when setting new clinical guidelines. This is especially important when facing the increasing variety of new, broad-spectrum antibiotics [22].

In this study, we present a test case which compares antibiotics commonly used in the community setting and for community-acquired infections among hospitalized patients. Our results are relevant for two clinical decision points. First, the need to choose an empirical therapy. Second, after bacteriology resistance tests are available, enabling a switch to more appropriate, definitive therapy.

This study had several limitations. First, the results of the comparison might not be relevant for various other clinical scenarios. Nosocomial infections, infections in immunocompromised or ICU patients might need assessment of broader spectrum antibiotics. The choice of a preferred empiric regimen for use in such scenarios is critical due to the complexity of these patient populations and their increased risk for future recurrent and resistant infections.

Moreover, the outcome used in this work — resistance profile to ceftazidime in a future culture within a year — is based on several prerequisites. The cohort examined here required the patients to survive, get infected, be hospitalized and tested for ceftazidime resistance. Hence, we implicitly assumed that replacing amoxicillin/clavulanate or cefazolin with cefuroxime would not have substantially affected these events. Another limitation is that our outcome is estimated only on rehospitalization. The NNH for patients who did not return to receive the outcome may prove different, and the estimated effect itself might have a different interpretation [23]. Obtaining an effect estimate on these patients would require data on culture results taken in the community or through active surveillance.

As with any observational study, unmeasured confounding could have biased our results. For example, we did not have information on antibiotic exposure in the community, which could influence the probability of acquisition of resistance if common and differentially distributed among the examined treatment groups. However, we chose antibiotics comparable to those prescribed for community-acquired infections; thus, community healthcare use is not expected to be fundamentally different. Future work could address the potential biases mentioned above by incorporating data from community usage and addressing the potential selection bias by methods such as inverse probability of censoring weighting [17].

Finally, our results are based on data from a single medical center. Other settings, including different resistance rates and antibiotic use patterns might lead to different resistance acquisition probabilities; thus, limiting the generalizability of the results.

In conclusion, we present an application of a statistical method which allows quantification of the ecological damage of different antibiotics. Further research is needed in different settings and with different antibiotics. This information will allow for a more knowledgeable use of antibiotics, while preserving future antibiotic effectiveness.

## Data Availability

Data produced in the present study are proprietary but can be available upon reasonable request to the authors

